# MR Imaging Distinguishes Tumor Hypoxia Levels of Different Prognostic and Biological Significance in Cervical Cancer

**DOI:** 10.1101/2020.05.28.20114769

**Authors:** Tiril Hillestad, Tord Hompland, Christina S. Fjeldbo, Vilde E. Skingen, Unn Beate Salberg, Eva-Katrine Aarnes, Anja Nilsen, Kjersti V. Lund, Tina S. Evensen, Gunnar B. Kristensen, Trond Stokke, Heidi Lyng

## Abstract

Tumor hypoxia levels range from mild to severe and have different biological and therapeutical consequences, but are not easily assessable in patients. We present a method based on diagnostic dynamic contrast enhanced (DCE) magnetic resonance imaging (MRI) that visualizes a continuous range of hypoxia levels in tumors of cervical cancer patients. Hypoxia images were generated using an established approach based on pixel-wise combination of the DCE-MRI parameters *ν*_e_ and *K*^trans^, reflecting oxygen consumption and supply, respectively. An algorithm to retrieve hypoxia levels from the images was developed and validated in 28 xenograft tumors, by comparing the MRI-defined levels with hypoxia levels derived from pimonidazole stained histological sections. We further established an indicator of hypoxia levels in patient tumors based on expression of nine hypoxia responsive genes. A strong correlation was found between these indicator values and the MRI-defined hypoxia levels in 63 patients. Chemoradiotherapy outcome of 74 patients was most strongly predicted by moderate hypoxia levels, whereas more severe or milder levels were less predictive. By combining gene expression profiles and MRI-defined hypoxia levels in cancer hallmark analysis, we identified a distribution of levels associated with each hallmark; oxidative phosphorylation and G_2_/M checkpoint were associated with moderate hypoxia, and epithelial-to-mesenchymal transition and inflammatory responses with significantly more severe levels. At the mildest levels, interferon response hallmarks, together with stabilization of HIF1A protein by immunohistochemistry, appearred significant. Thus, our method visualizes the distribution of hypoxia levels within patient tumors and has potential to distinguish levels of different prognostic and biological significance.

## Introduction

Solid tumors show a highly heterogeneous oxygen distribution with hypoxia levels ranging from mild to moderate and severe (1). The hypoxia level determines resistance to cancer therapies like radiation, chemotherapy and many molecular targeting drugs (1–4), and may therefore have large therapeutical consequences. Current understanding of how the different levels drive cancer progression and affect treatment response is scarce and mostly based on experimental studies (5–7). At mild hypoxia, around 2% O_2_, activation of the hypoxia inducible transcription factor HIF1 promotes metabolic reprogramming and cell survival (8,9), more severe levels, below 1% O_2_, may impair cell proliferation and lead to genomic instability (10,11), and below 0.5% O_2_ the cytotoxic effect of radiation is more than 2-fold reduced (2). Hypoxia may also induce epithelial-mesenchymal-transition (EMT) and immune evasion of tumor cells (12,13), but the levels of importance for these processes have not been clarified. In patient tumors, earlier investigations using invasive electrodes to measure oxygen partial pressure (pO_2_) have shown considerable differences across cancer types in the level most strongly associated with treatment outcome, ranging from 2.5-10 mmHg or approximately 0.3-1.3% O_2_ (14). More recent clinical work has almost exclusively focused on the presence or absence of hypoxia (15), mainly because oxygen electrodes are not feasible and alternative approaches to assess hypoxia levels are lacking. A method based on medical imaging would facilitate investigations of how individual levels relate to treatment outcome and tumor biology in patients, and help development of more efficient therapies to combat hypoxia.

Hypoxia occurs in tumors due to impaired oxygen supply by a chaotic vascular network and/or elevated oxygen consumption in regions with high cellularity (1). We recently presented a tool for pixel-wise combination of images reflecting oxygen consumption with images reflecting oxygen supply into images representing hypoxia (16). The consumption and supply based hypoxia (CSH)-imaging tool was originally developed in prostate cancer patients, using images of the apparent diffusion coefficient (ADC) and fractional blood volume (fBV) derived from diffusion weighted (DW) magnetic resonance (MR) images. The information in the two images was utilized to reflect the difference between oxygen consumption and supply and thereby the probability of each pixel to locate in a hypoxic region. Although only the presence of hypoxia was addressed in this study, it is likely that a difference between oxygen consumption and supply within a tumor region also would provide information on the hypoxia level. The CSH-principle may therefore be a basis for establishing an imaging approach for quantifying hypoxia levels.

Locally advanced cervical cancer is a disease for which better biological understanding and new therapeutical approaches to overcome hypoxia are urgent (17,18). In the present work, we aimed to construct images that visualize a continuous distribution of hypoxia levels in cervical tumors by applying the CSH-tool. Our approach was based on dynamic contrast enhanced (DCE)-MR imaging (MRI), because this modality is state-of-the-art diagnostics for the disease. We showed that the DCE-MRI parameters *ν*_e_ and *K*^trans^ from the Tofts pharmacokinetic model (19) reflected oxygen consumption and supply, respectively, and could be successfully combined to generate hypoxia images in xenograft and patient tumors. We further developed an algorithm to assign hypoxia levels to all pixels. The algorithm was validated by comparison with hypoxia levels determined from pimonidazole stained sections in xenograft tumors and hypoxia related gene expression in patient tumors. The power of this approach was demonstrated by presenting the distribution of hypoxia levels in tumors of 74 patients and identifying significant differences in the levels associated with treatment outcome and a set of cancer hallmarks.

## Materials and Methods

### Clinical cohort

Totally 74 patients with locally advanced cervical carcinoma, prospectively recruited to our chemoradiotherapy protocol at the Norwegian Radium Hospital were included (Supplementary Table S1). Gene expression profiles and a gene score reflecting hypoxia were available from previous work (20) for 63 patients, and paraffin embedded tissue sections for immunohistochemistry were available for 73 patients. The gene score was based on the expression level of 6 hypoxia responsive genes and increased with increasing amount of hypoxia (20). All patients received external radiotherapy combined with cisplatin (40 mg/m^2^ weekly) followed by intracavitary brachytherapy and follow up as described (20). The study was approved by the Regional Committee for Medical Health Research Ethics in southern Norway, and written informed consent was attained from all patients.

### Cell lines and hypoxia treatment

HeLa and SiHa cervical cancer cell lines from American Type Culture Collection were used. Confirmation of cell line identity and cell culturing were performed as described (21). Totally 1.5·10^6^ HeLa and 1.7·10^6^ SiHa cells were reseeded in 10 cm plastic dishes 24 hours before exposure to hypoxia at 0.2%, 0.5%, 1%, 2% and 5% O_2_ for 24 hours at 37ºC, all with 5% CO_2_, by using an Invivo_2_200 chamber (Ruskinn Technology Ltd). Normoxic controls (95% air, 5% CO_2_) were included for all hypoxia experiments.

### Human tumor xenografts

HeLa and SiHa cervical cancer xenograft tumors were established in female nude mice, bred at the animal department of our institute and kept in specific pathogen-free environment, with food and water supplied ad libitum. Totally 1·10^6^ HeLa cells in 20 μl or 2·10^6^ SiHa cells in 40 μl of Hank’s balanced salt solution were injected intramuscularly in both hind legs of adult mice. Tumor growth was monitored with anatomical T_2_-weighted MRI. At the day of DCE-MRI, the hypoxia marker pimonidazole (60 mg/kg; Hydroxyprobe, Inc) was administered intraperitoneally prior to MR scanning in 16 HeLa and 12 SiHa tumors. After the scan, 90-120 minutes after pimonidazole injection, the mice were euthanized by dislocation of the neck, and the tumors were excised, formalin-fixed and paraffin-embedded for immunohistochemistry. All procedures were approved by the Norwegian Animal Research Authority and performed in accordance with the guidelines on animal welfare of the Federation of Laboratory Animal Science Associations.

### DCE-MRI

DCE-MRI of xenograft tumors was performed at a volume of 100-800 mm^3^, using a 7.05 T Biospec bore magnet (Bruker) and a fast bolus injection of 5.0 ml/kg body weight of Gd-DOTA (Dotarem, Guerbet) (Supplementary Method S1). Totally 8 images prior to and 57 images post injection of Gd-DOTA were acquired with an axial T_1_-weighted spoiled gradient recalled sequence (SPGR). The images had a spatial resolution of 234×234×1000 μm^3^. The three most central tumor slices were used in the analysis.

In patients, DCE-MRI was performed at diagnosis, using a 1.5 T Signa Horizon LX tomograph (GE Medical Systems) with a pelvic phased array coil and a fast bolus injection of 0.1 mmol/kg body weight Gd-DTPA (Magnevist, Schering) (Supplementary Method S1). Totally 1-2 series prior to and 12-13 series post injection of Gd-DTPA were acquired with an axial T_1_-weighted SPGR sequence. The images had a pixel size of 780×780 μm^2^, slice thickness of 5 mm and slice gap of 1 mm. All slices containing tumor were used in the analysis.

### Hypoxia images

The tumors were outlined in T2-weigthed MR images and co-registered with the DCE-MR images. Pharmacokinetic analysis of the contrast uptake curves obtained from the DCE-MR images was performed on a pixel-by-pixel basis using the Tofts model (19) (Supplementary Methods S1), and parametric images of *K*^trans^ and *ν*_e_ were generated. To construct hypoxia images, the CSH-tool was applied on the *K*^trans^ and *ν*_e_ images as described for DW-MRI (16). Hence, pixel-wise plots of *K*^trans^ *versus ν*_e_ were generated for each tumor, representing decreasing oxygen consumption on the horizontal *ν*_e_-axis and increasing oxygen supply on the vertical *K*^trans^-axis. To determine a threshold for hypoxia, a line discriminating pixels in hypoxic and non-hypoxic regions, and thus defining the hypoxic fraction, was determined in an iterative procedure with all tumors, using an independent hypoxia measure as learning variable. The hypoxic fraction was calculated for each tumor and iteration and correlated with the independent hypoxia measure. The optimal line was determined by the highest Pearson correlation coefficient and was described by its intersections with the horizontal (*ν*_e0_) and vertical axes (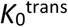).

### Immunohistochemistry and digital histopathology

Adjacent sections, 4-5 μm thick, from xenograft tumors were stained for hypoxia (n=28) using a pimonidazole polyclonal rabbit antibody (1:3500; Hydroxyprobe Inc.) and endothelial cells (n=26), using a CD31 rabbit polyclonal antibody (1:50, ab28364; Abcam). Hematoxylin was used as counterstain to visualize cell nuclei. Digital histopathology was performed to quantify hypoxic fraction (*HF*_Pimo_), cell density (CD) and blood vessel density (BVD) (Supplementary Method S1). Sections from 73 patient tumors were stained with the monoclonal mouse HIF1A antibody clone 54 (1:25, no. 610958; BD Transduction Laboratories) as described (21). Percentage of HIF1A positive tumor cells was scored manually based on nuclear staining: 0, 0%; 1, 1-10%; 2, 11-25%; 3, 26-50%; 4, 51-75% and 5, >75%.

### Gene expression

Gene expression profiling of HeLa and SiHa cells exposed to hypoxia at 0.2%, 0.5%, 1%, 2% and 5% O_2_ and normoxia (95% air) was carried out using Illumina bead arrays HT-12 v4 (Illumina Inc.). Total RNA was isolated using miRNeasy MiniKit (Qiagen). Complementary RNA was synthesized, labeled and hybridized to the arrays. Signal extraction and quantile normalization were performed using software provided by the manufacturer (Illumina Inc.). The data were deposited in the Gene Expression Omnibus (GEO; GSE147384). Normalized gene expression profiles of 63 patients, generated previously using Illumina bead arrays WG-6 v3 (Illumina Inc.) (20), were downloaded from GEO (GSE72723).

### Statistical analysis

To compare hypoxic fractions derived from MR images and pimonidazole stained sections, an adapted version of Pearson product moment correlation test for similarity between two data sets was applied (22):

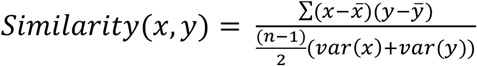

where *x* and *y* are sets of hypoxic fractions from MRI and pimonidazole, respectively, *var*(*x*) and *var*(*y*) are their sample variance and *n* is sample size. The function is equal to one when the hypoxic fractions from the two modalities are perfectly correlated with a slope of one. In cases of poor correlation or a slope deviating from one, the similarity decreases.

Curve fitting was performed through regression analysis. Student’s t-test was used for comparison of groups when data complied conditions of normality and equal variance. Otherwise, Wilcoxon rank sum test was used. Linear correlations were searched for by Pearson correlation test. Clinical endpoint was progression-free survival defined as time from diagnosis to disease-related death or first occurrence of relapse. Patients were censored at their last appointment or at 5 years. Cox univariate proportional hazard analysis was performed, and Kaplan-Meier curves were compared using log-rank test. Probability values of *P*<0.05 were considered significant. The statistical analyses were performed using SigmaPlot and SPSS.

## Results

### MRI-based hypoxia images provide measures of hypoxic fraction

The possibility to construct hypoxia images from DCE-MR images was investigated in xenograft tumors by first examining whether the histopathology parameters cell density (CD) and blood vessel density (BVD) could be used to reflect oxygen consumption and supply, respectively. Images of the DCE-MRI parameters *ν*_e_ and *K*^trans^ displayed resemblance with those of CD and BVD, respectively, with some disagreement possibly due to a two-hundred fold difference in slice thickness (Supplementary Fig. S1). Consistent with these observations, significant correlations were found between median values of *ν*_e_ and CD (*R*^2^=0.46, *P*<0.0005) and between median values of *K*^trans^ and BVD (*R*^2^=0.17, *P*=0.03) (Supplementary Fig. S1). No significant correlation was found between *ν*_e_ and BVD or between *K*^trans^ and CD (Supplementary Fig. S2A). Hypoxic fraction determined by pimonidazole staining (*HF*_Pimo_) was correlated with both *ν*_e_ (*R*^2^=0.46, *P*<0.00005) and *K*^trans^ (*R*^2^=0.22, *P*<0.05; Supplementary Fig. S2B). *ν*_e_ and *K*^trans^ therefore seemed to be connected to hypoxia and contain different information related to oxygen consumption and supply, respectively, in line with other reports where low molecular weight contrast agents are used for DCE-MRI (23).

Based on the above results, we searched to construct hypoxia images in xenograft tumors by combining images of *ν*_e_ and *K*^trans^ and using *HF*_Pimo_ as independent measure of hypoxia. In pixel-wise plots of *K*^trans^ *versus ν*_e_, pixels from tumors having a high *HF*_Pimo_ were in general located more towards the lower left corner than pixels from tumors with a low *HF*_Pimo_ (Fig. 1A, B), consistent with the CSH-principle. The line that best discriminated pixels in hypoxic and non-hypoxic regions for all tumors combined was determined (Supplementary Fig. S3A). Pixels below the optimal line were considered hypoxic and the fraction of these pixels, *HF*_MRI_, was strongly correlated to *HF*_Pimo_ (*R*^2^=0.57, *P*<0.000005; Fig. 1C). This correlation was stronger than between *ν*_e_ or *K*^trans^ and *HF*_Pimo_ (Supplementary Fig. S2). The resulting binary hypoxia images showed strong resemblance to the pimonidazole stained sections (Fig. 1B, D).

**Figure 1.**
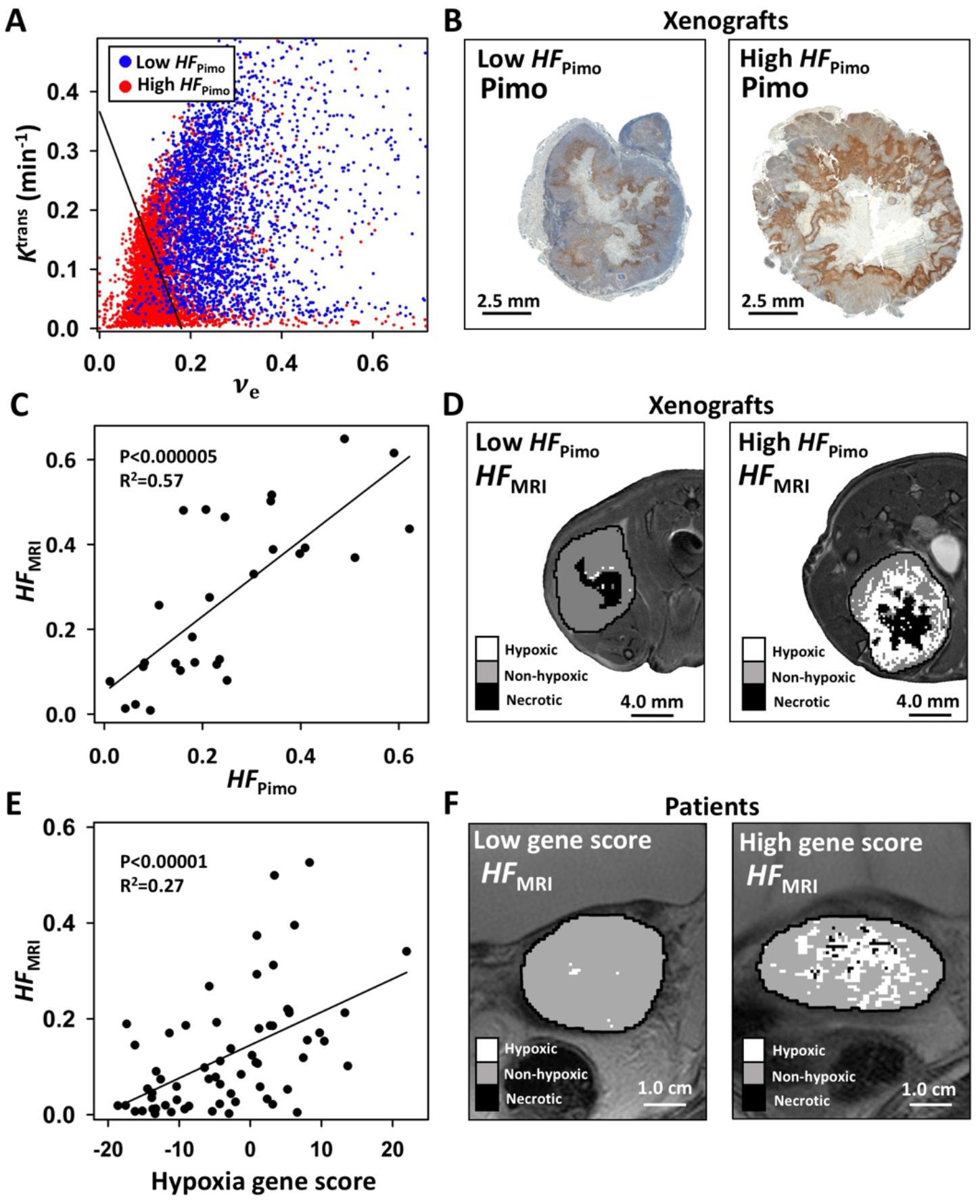
Construction of hypoxia images in xenograft and patient tumors. **A**, Pixel-wise plot of *K*^trans^ *versus v*_e_ for a xenograft tumor with high hypoxic fraction according to pimonidazole staining (*HF*_Pimo_) (red) and another with low *HF*_Pimo_ (blue). The optimal discrimination line separating pixels in hypoxic and non-hypoxic regions is shown. **B**, Pimonidazole stained sections of the tumors presented in **A**. **C**, Scatter plot of *HF*_MRI_ *versus HF*_Pimo_ for 28 xenograft tumors based on the optimal discrimination line. **D**, Binary hypoxia images visualizing *HF*_MRI_ of the tumors presented in **A** and **B**. **E**, Scatter plot of HF_MRI_ *versus* hypoxia gene score for 63 patient tumors based on the optimal discrimination line. **F**, Binary hypoxia images visualizing *HF*_MRI_ of a less and more hypoxic tumor according to the hypoxia gene score. **C**, **E**, *P*-value and correlation coefficient (*R^2^*) from linear correlation analysis are shown. **D**, **F**, The binary images are overlaid on axial T_2_-weighted images.

To confirm applicability of the CSH-tool to produce hypoxia images in patient tumors, pixel-wise plots of *K*^trans^ *versus ν*_e_ were generated from the clinical images. Similar to what we observed in xenografts, pixels from hypoxic tumors appeared to be located towards the lower left corner in these plots (Supplementary Fig. S4). By using the same procedure as above and the gene score from previous work (20) as independent hypoxia measure, an optimal line to discriminate pixels in hypoxic and non-hypoxic regions for all tumors combined was determined (Supplementary Fig. S3B) and a *HF*_MRI_ was calculated for each tumor. A strong correlation between *HF*_MRI_ and the hypoxia gene score (*R*^2^=0.27, *P*<0.00001; Fig. 1E, F) was found. This correlation was stronger than between *ν*_e_ or *K*^trans^ and the gene score (Supplementary Fig. S5A). In analysis of all 74 patients, *HF*_MRI_ was strongly correlated with progression-free survival, where patients with high *HF*_MRI_ had a poor outcome compared to the others (*P*=0.0014; Supplementary Fig. S5B), consistent with the prognostic significance of the gene score (20). The correlation to outcome was weaker for *K*^trans^ or *ν*_e_ (*P*=0.015 and *P*=0.074, respectively; Supplementary Fig. S5B). All together, this showed that hypoxia images could be constructed using the DCE-MRI parameters *ν*_e_ and *K*^trans^ as input to the CSH-tool.

### Hypoxia levels defined by pimonidazole staining in xenograft tumors are visualized by MRI

Based on the hypoxia images, an algorithm to assign a hypoxia level to each individual pixel was developed. We hypothesized that the location of a pixel in plots of *K*^trans^ *versus ν*_e_; *i.e*., the distance from the pixel to the optimal discrimination line, depends on the hypoxia level of the corresponding tumor region (Fig. 2A). This hypothesis is likely because the line represents the weighted information of *K*^trans^ (oxygen supply) and *ν*_e_ (oxygen consumption) underlying the level of the independent hypoxia measure. The hypoxia level, *HL*_MRI_, can thus be expressed as:

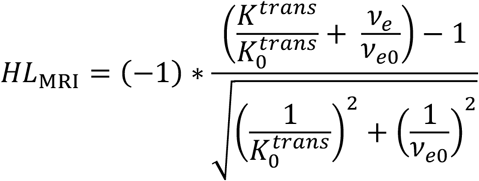

where the level of the optimal line, described by the intersection points *ν*_e0_ and 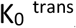, was set to zero, and increasing values of *HL*_MRI_ indicated more severe hypoxia. Application of the algorithm to calculate four hypoxia levels is shown in Figure 2B, together with the underlying *HL*_MRI_ image (Fig. 2C).

**Figure 2.**
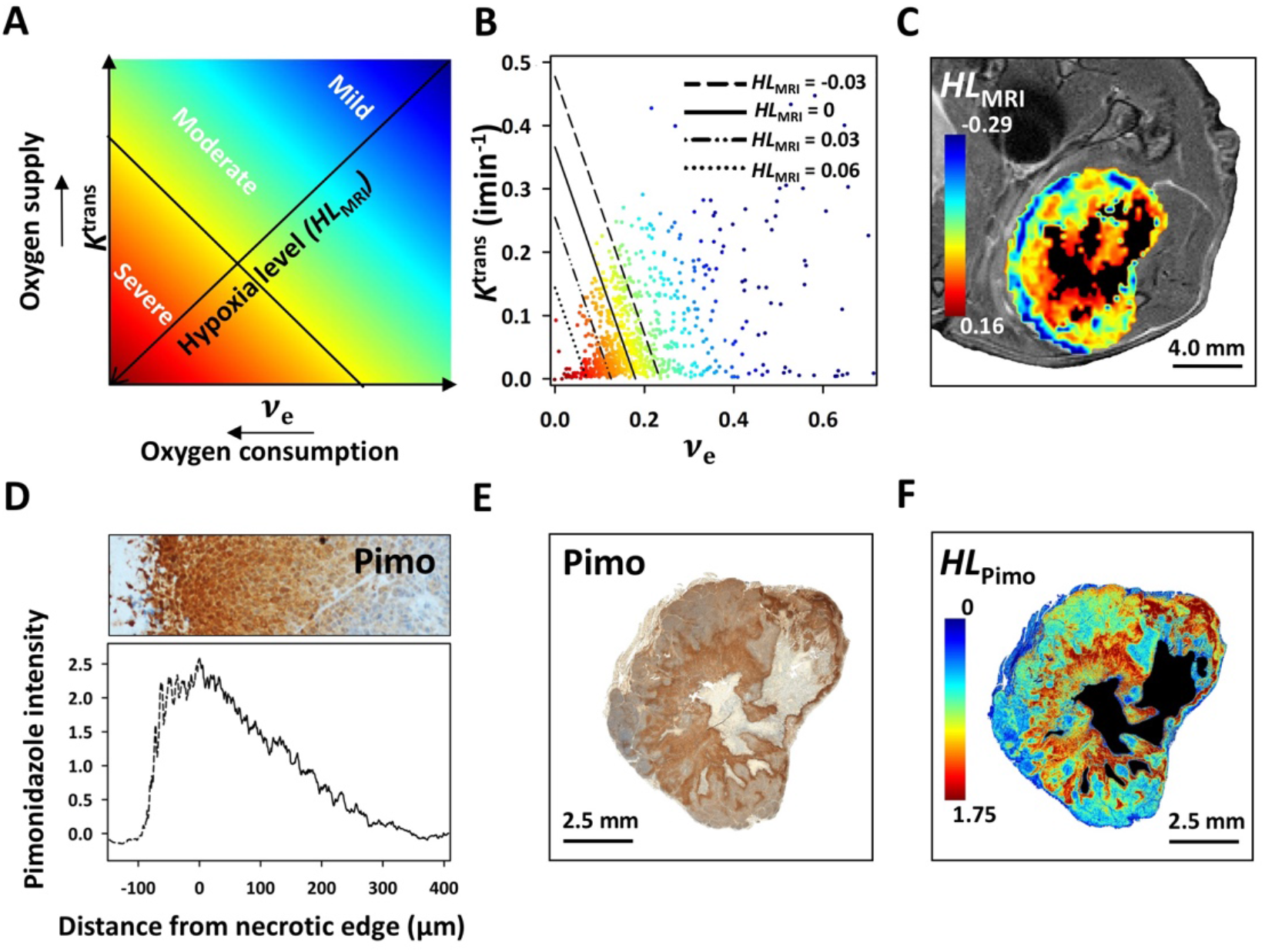
Indicator of hypoxia levels in xenograft tumors. **A**, Principle of assessing hypoxia levels (*HL*_MRI_) from hypoxia images as the distance from the pixel to the optimal discrimination line (*HL*_MRI_=0). **B**, Pixel-wise plot of *K*^trans^ *versus v*_e_ of a xenograft tumor. The solid line indicates the optimal discrimination line (*HL*_MRI_=0), whereas the stippled lines in parallel represent three different hypoxia levels; i.e. different *HL*_MRI_ values. Points are color-coded according to their *HL*_MRI_ value. **C**, Hypoxia image of the tumor presented in **B**, overlaid on an axial T_2_-weighted image. **D**, Pimonidazole staining intensity in histological sections from a xenograft tumor *versus* distance from necrosis. The histological section is shown above. **E**, Pimonidazole stained section of the tumor presented in **B** and **C**. **F**, Color coded pimonidazole-based image of hypoxia levels, *HLpj_mo_*, for the tumor presented in **B**, **C** and **E**.

A procedure to extract hypoxia levels from pimonidazole stained tumor sections in xenografts was developed for validation of the algorithm. *In vitro* studies have shown that the binding efficacy of pimonidazole during hypoxia increases exponentially with decreasing oxygen concentrations (24). In line with this, the pimonidazole staining intensity was generally strongest close to necrotic regions (anoxia) and decreased with increasing distance from necrosis (Fig. 2D), most likely reflecting a hypoxia gradient. We therefore assumed that the staining intensity was proportional to hypoxia level, and produced pimonidazole-based images of hypoxia levels (*HL*_Pimo_) that were used for validation (Fig. 2E, F; Supplementary Method S1). Visual inspection showed large resemblance between the *HL*_MRI_ and *HL*_Pimo_ images (Fig. 2C, F), although there was a considerable difference in slice thickness between the two modalities. By this inspection, we further found that the staining intensity in pimonidazole-based images could be evaluated down to a *HL*_Pimo_ of 0.38. Below this limit, the intensity was weak with small changes, probably reflecting non-hypoxic levels.

Hypoxia levels derived from MR- and pimonidazole-based images (Fig. 2C, F) were compared in 28 xenografts. By varying the threshold for *HL*_MRI_, from 0.11 in severe hypoxia to −0.05 at the mildest level, and for *HL*_Pimo_, from 2.6 at the strongest staining intensity to 0.01 in the weakly stained region, we generated sets of hypoxic fractions (% of tumor > *HL*_MRI_ or *HL*_Pimo_) for both modalities and all xenografts (Fig. 3A). The two data sets, each consisting of 28×200 hypoxic fractions, were first compared using similarity analysis, where we for each *HL*_MRI_ threshold identified the *HL*_pimo_ threshold that led to the highest similarity between hypoxic fraction derived by the two modalities (Fig. 3B). Overall, the similarity values were high (>0.6) and an exponential relationship was observed between the similarity-matched *HL*_MRI_ and *HL*_Pimo_. The exponential relationship, presented as a linear relationship in a log plot in Figure 3C, is in line with the exponential binding of pimonidazole with decreasing oxygen concentrations (24). Correlation analysis of the most similar hypoxic fractions provided an indication of how well *HL*_MRI_ reflected the different hypoxia levels. A strong correlation (*P*<0.001) was found for *HL*_MRI_ in the range of −0.03 to 0.1. Hence, hypoxic fraction from a large range of levels could be measured. Moreover, within this range the mean hypoxic fraction based on all 28 xenograft tumors showed considerable differences, ranging from 0.38 at mild hypoxia (*HL*_MRI_=-0.03; Fig. 3D) to 0.07 at more severe hypoxia (*HL*_MRI_=0.06; Fig. 3E) and 0.02 at the most severe levels (*HL*_MRI_=0.1; data not shown). These results supported that our algorithm to image hypoxia levels was reliable. Further, the MRI-defined hypoxia levels could distinguish a large range of hypoxic fractions in xenograft tumors.

**Figure 3.**
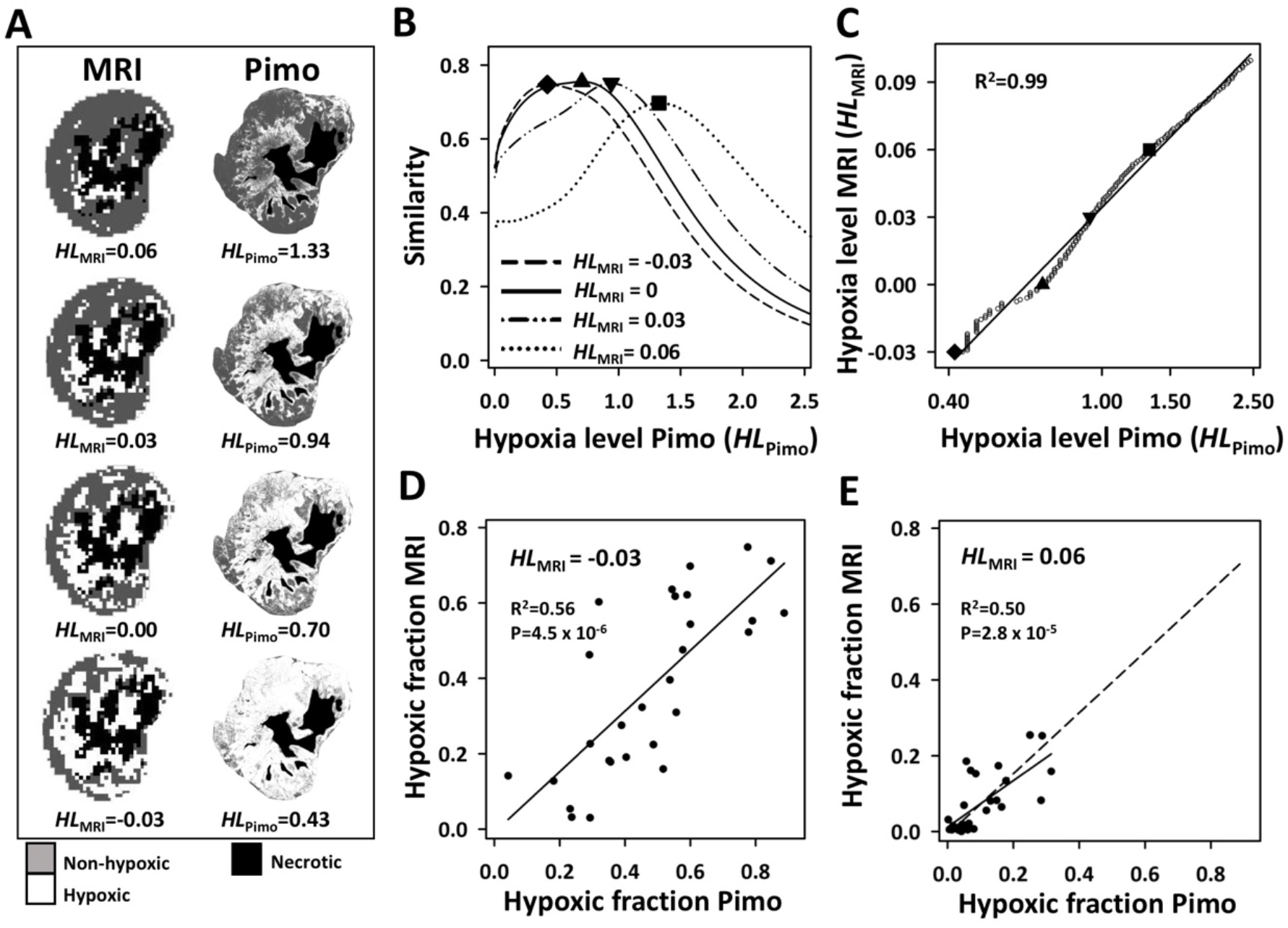
Assessing hypoxia levels in xenograft tumors. **A** Examples of binary MR- and pimonidazole-based images, visualizing hypoxic fractions for four different *HL*_MRI_ and *HL*_Pimo_ thresholds of the tumor presented in **Fig. 2C** and **2F**. **B**, Similarity plots for the *H*_MRI_ thresholds indicated in **A**, showing the similarity between MRI- and pimonidazole-based hypoxic fractions *versus H*_Pimo_ threshold. The highest similarity is marked for each *H*_MRI_ threshold. **C**, *H*_MRI_ *versus H*_Pimo_ in 28 xenograft tumors. Similarity-matched *H*_MRI_ and *H*_Pimo_; i.e., *HL*_MRI_ and *HL*_Pimo_ leading to the highest similarity in the analysis presented in **B**, are shown. The four *HL*_MRI_ thresholds shown in A are indicated with solid symbols together with the correlation coefficient (*R^2^*) and curve from linear correlation analysis. **D, E**, Scatterplots of MRI-based versus pimonidazole-based hypoxic fraction for a *HL*_MRI_ threshold of −0.03 (**D**) and 0.06 (**E**). Similarity-matched *HL*_MRI_ and *HL*_Pimo_ were used to calculate hypoxic fractions for 28 xenograft tumors. *P*-value, correlation coefficient (*R^2^*) and curve from linear correlation analysis are shown.

### Hypoxia levels defined by gene expression in patient tumors are visualized by MRI

To confirm the validity of our algorithm in patient tumors, we constructed an indicator of hypoxia levels based on the expression of hypoxia responsive genes. We utilized that genes may be activated and, thus, show increased expression, at specific oxygen concentrations(25). Nine indicator genes were selected among 31 previously identified hypoxia responsive genes in cervical cancer (21) (Supplementary Document S1). The genes are known to be regulated by HIF1 (*AK4, PFKFB4, P4HA2*), by both HIF1 and the unfolded protein response (*STC2, ERO1A*) or the regulation mechanisms are poorly exlpored (*UPK1A, KCTD11, SNTA1, PYGL*). By exposure of SiHa and HeLa cells to oxygen concentrations in the range of 0.2-21% O_2_, the concentration for half-maximal response was recorded for each gene (Fig. 4A, B), in a similar way as described for stabilization of HIF1A protein (8). This cell line derived hypoxia activation level was found to range from 0.55% to 1.81% O_2_, where the HIF1A targets *AK4* and *PFKFB4* had the highest level, in line with an HIF1 activation level of 1.5-2.0% O_2_ (8) (Fig. 4C, Supplementary Document S1). Thus, the indicator genes showed a range of levels likely to be found in human tumors (6) and broad enough for testing our algorithm.

**Figure 4.**
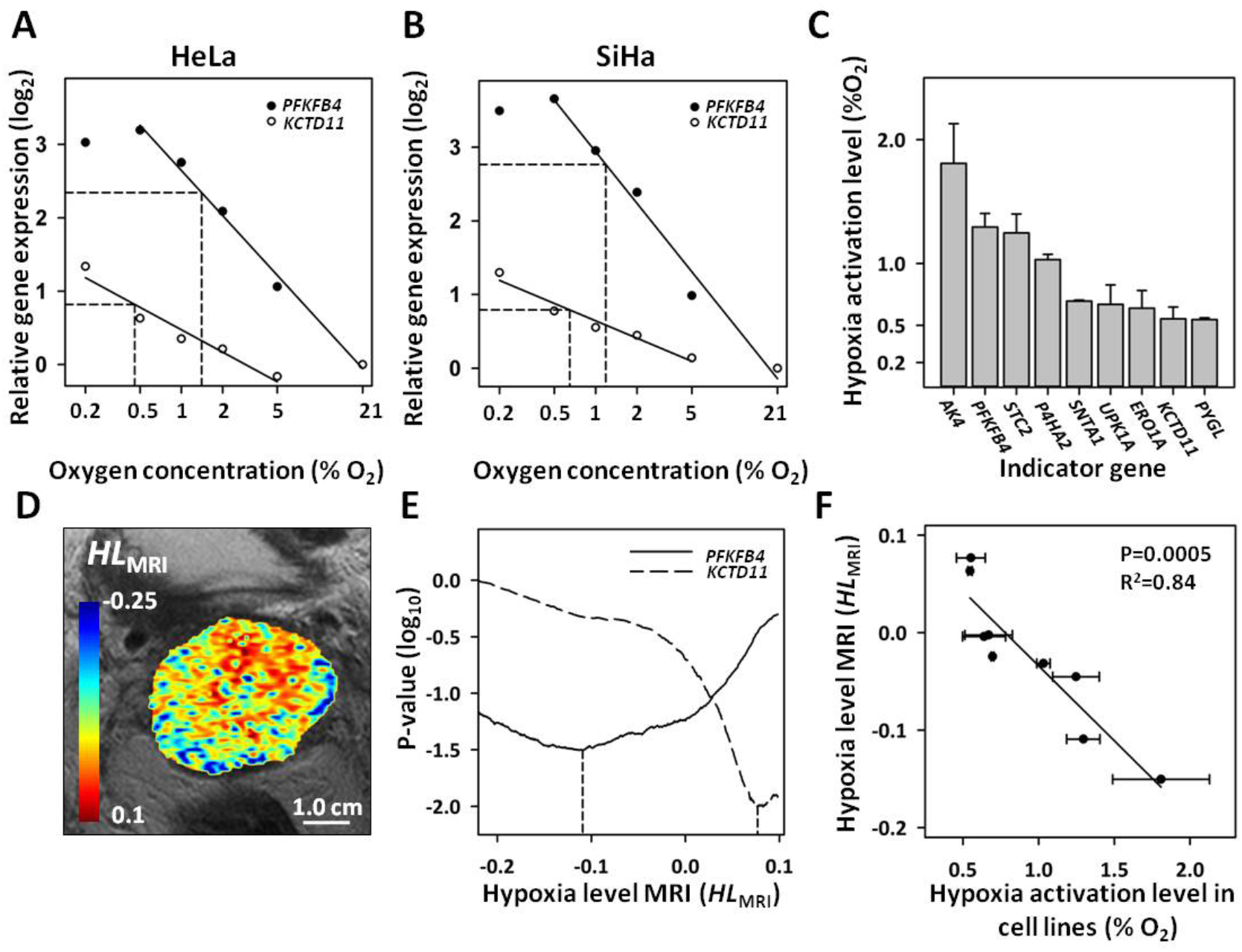
Assessing hypoxia levels in patient tumors. **A**, **B**, Gene expression in HeLa (**A**) and SiHa (**B**) cell lines *versus* the logarithm of oxygen concentration for two indicator genes, *KCTD11* and *PKFKB4*. The expression levels are plottet relative to the level of normoxic controls (21% O_2_). Hypoxia activation level and curve from linear correlation analysis are indicated for each gene. **C**, Hypoxia activation level of nine indicator genes. Bars, range of data for SiHa and HeLa cell line. **D**, Hypoxia level image of a patient tumor overlaid on an axial T_2_-weighted image. **E**, P-value in correlation analysis of hypoxic fraction calculated for a set of 200 *HL*_MRI_ thresholds *versus* gene expression in 63 patients, plotted as a function of *HL*_MRI_. Data for two indicator genes, *KCTD11* and *PKFKB4* are shown. The *HL*_MRI_ value leading to the strongest correlation between gene expression and MRI-based hypoxic fraction (*i.e*., lowest P-value) is indicated for each gene. F, *HL*_MRI_ for the strongest correlation achieved in E *versus* hypoxia gene activation level in cell lines for nine indicator genes. Point and bar, average value and range for SiHa and HeLa cell lines. Curve, *P*-value and correlation coefficient (*R*^2^) from linear correlation analysis are shown.

*HL*_MRI_ images were constructed for all 74 patient tumors (Fig. 4D). Using the same strategy as for xenografts, a set of 200 hypoxic fractions was calculated for each tumor using *HL*_MRI_ thresholds ranging from 0.1 in severe hypoxia to −0.3 as the mildest level. Expression data of the nine indicator genes were further retrieved from the gene expression profiles of each tumor. A correlation analysis of the two data sets was performed, where we for each indicator gene identified the *HL*_MRI_ threshold that led to the strongest association between hypoxic fraction and expression (Fig. 4E; Supplementary Document S1). These *HL*_MRI_ thresholds showed a strong correlation to the cell line derived hypoxia activation level for the nine indicator genes (Fig. 4F; *R*^2^=0.84, *P*<0.0005). Although oxygen concentrations found for half-maximal response in cell lines are not directly transferable to patient tumors, this relationship together with the above xenograft results strongly supported that *HL*_MRI_ provided a continuous, linear measure of hypoxia levels in tumors.

### Hypoxia levels of prognostic significance are distinguished in MR images

The relationship presented in Figure 4F provided a tool to relate MRI defined hypoxia levels to biological information derived in cell lines. Aided by this relationship, we defined approximate *HL*_MRI_ intervals for severe, moderate and mild hypoxia in order to characterize the hypoxia level distribution in patient tumors (Fig. 5A). The definitions corresponded roughly to those proposed by others (6). Median *HL*_MRI_ of all tumors combined was −0.08. This value was related to a cell line derived level of 1.3% O_2_ (Fig. 5A) and within the moderate hypoxia range. However, the median value differed considerably across tumors, ranging from −0.22 (2.3% O_2_) in mild hypoxia to 0.004 (0.8% O_2_) in moderate hypoxia. A pie chart of each tumor was generated to visualize these differences, showing fraction of pixels within *HL*_MRI_ intervals of 0.05 (Fig. 5B; Supplementary Fig. S6). Most tumors contained a range from severe to non-hypoxic levels, however, fraction of the different levels varied considerably across patients.

**Figure 5.**
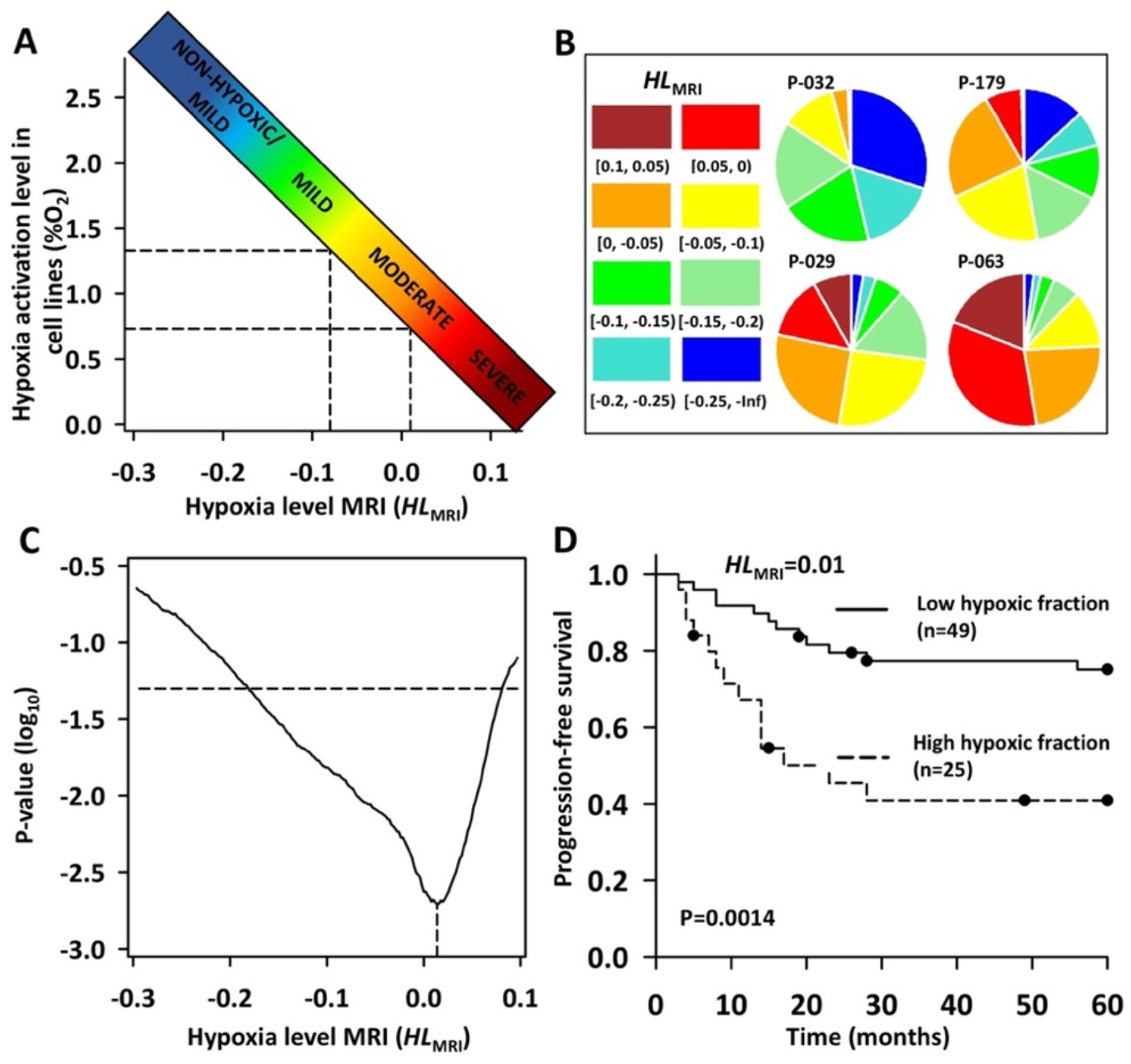
Hypoxia levels in patient tumors in relation to treatment outcome. **A**, Approximate *HL*_MRI_ intervals for severe, moderate and mild hypoxia based the relationship between *HL*_MRI_ and hypoxia gene activation level in cell lines presented in **Fig. 4F**. Stippled lines indicate median hypoxia level (*HL*_MRI_=-0.08) for all patient tumors combined and the hypoxia level with the strongest correlation to progession free survival (*HL*_MRI_=0.01) in the analysis presented in **C. B**, Pie charts showing fractions of pixels with *HL*_MRI_ within the indicated intervals for four tumors with different distribution of hypoxia levels. **C**, *P*-value in Cox regression analysis of hypoxic fraction calculated for increasing *HL*_MRI_ threshold (increasing severity level) *versus* progression-free suvival, plotted as a function of *HL*_MRI_. Horizontal stippled line indicates a significance level of 0.05. Vertical stippled line indicates *HL*_MRI_ for the strongest correlation (*HL*_MRI_=0.01). **D**, Kaplan Meier curves for progression-free survival of 74 patients with low (solid line) and high (stippled line) hypoxic fraction based on the *HL*_MRI_ threshold of 0.01 indicated in **C**. Patients were divided with 1/3 in the high-risk and 2/3 in the low-risk group based on an expected failure rate 30%. *P*-value in log-rank test is shown.

To address whether differences seen in the pie charts across patients were associated with differences in chemoradiotherapy outcome, hypoxic fraction was determined for a range of *HL*_MRI_ thresholds for each tumor and included in survival analysis with progression-free survival as end point. The strongest association to outcome was found for hypoxic fractions below a *HL*_MRI_ threshold of 0.01 (Fig. 5C), which was related to a cell line derived level of 0.7% O_2_ and in moderate hypoxia close to the interval of severe hypoxia (Fig. 5A). Hence, patients with a high hypoxic fraction below this level had a poor outcome compared to the others (P=0.0014; Fig. 5D). In contrast, weaker or no association to outcome was found for more severe hypoxia; *i.e*., the highest *HL*_MRI_ values, or for milder hypoxia.

### MR images distinguish hypoxia levels of biological significance

The data set of hypoxic fractions generated in the above analysis was further correlated with gene expression profiles of the patient tumors to identify possible associations between hypoxia levels and biological processes. Totally 1344 genes showed a positive correlation (*P*<0.05) for one or more *HL*_MRI_ thresholds and were included in a hallmark enrichment analysis. Out of 50 hallmarks, 36 were found to be significantly enriched (Supplementary Table S2), and 350 of the 1344 genes were included in one or more of these hallmarks. By assigning the *HL*_MRI_ threshold showing the strongest correlation between hypoxic fraction and expression (P<0.05) for the 350 genes, a distribution of hypoxia levels was produced for each of the 36 enriched hallmarks. In general, the individual *HL*_MRI_ distributions covered a large range of hypoxia levels, and most hallmarks (n=26) had a median *HL*_MRI_ in the moderate hypoxia range, including well known hypoxia regulated processes like hypoxia and glycolysis (Supplementary Figure S7, S8).

The *HL*_MRI_ distributions were further compared across the 36 hallmarks, to search for differences in the hypoxia level associated with biological processes. All hallmarks were tested against each other, and those with a difference (*P*<0.05) to less than 25% of the others were removed to simplify analysis. For the remaining 15 hallmarks, three groups with a significant difference in *HL*_MRI_ distribution was identified (Fig. 6A; Supplementary Figure S7). A group with the interferon *α* and *γ* response hallmarks was associated with mild hypoxia (Fig. 6A, B). At moderate levels, a group including G_2_/M checkpoint, MYC targets, oxidative phosphorylation and MTORC1 signalling appeared significant, whereas hallmarks like TNFA signalling via NFKB, DNA repair, inflammatory response, angiogenesis and EMT were associated with the most severe levels.

**Figure 6.**
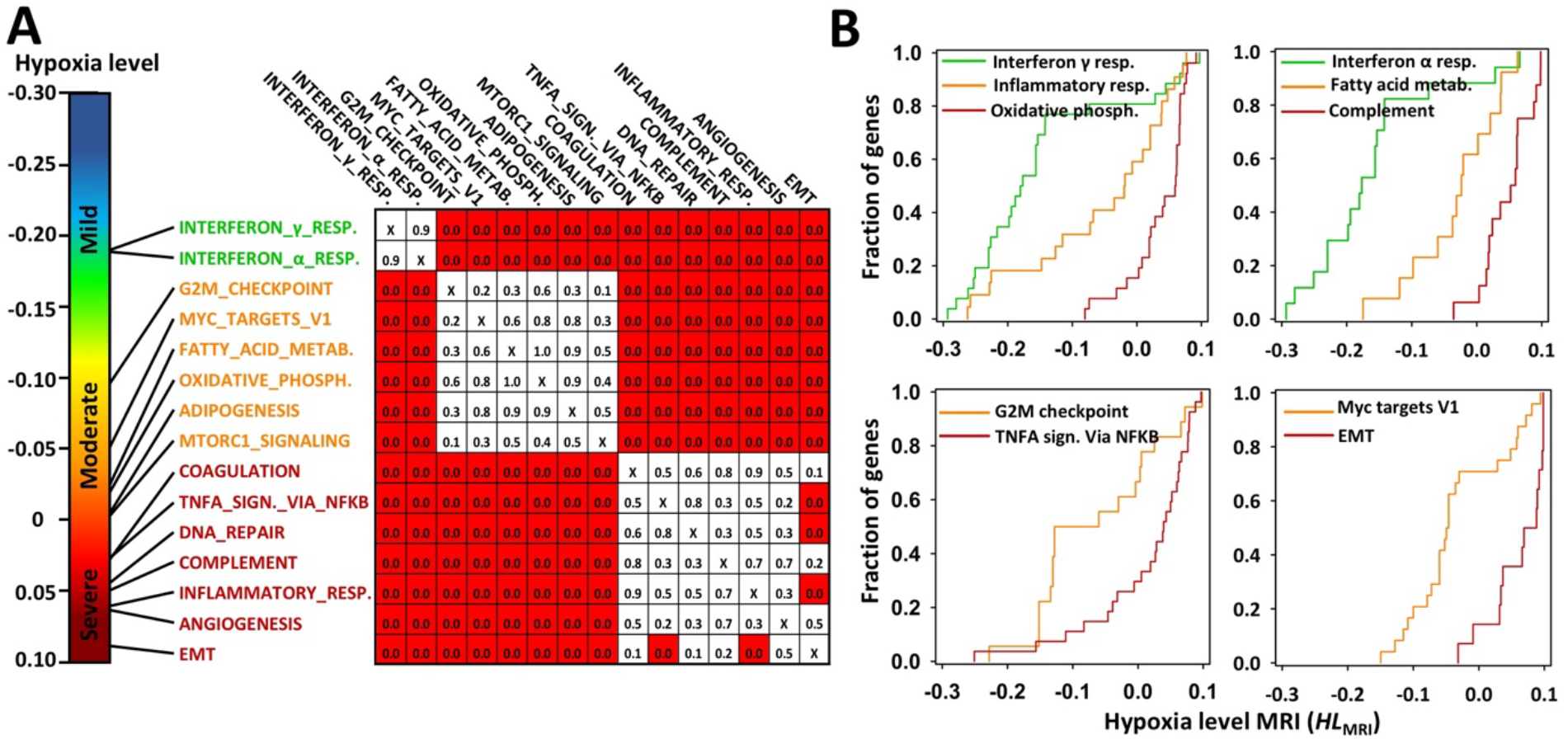
Hypoxia levels in patient tumors in relation to cancer hallmarks. **A**, Correlation analysis showing three distinct groups of hallmarks with significant difference in *HL*_MRI_ distribution, each related to either mild, moderate or severe hypoxia. *P*-values from Wilcoxon rank sum test are shown (right). **B**, Cumulative *HL*_MRI_ distribution associated with a selection of the hallmarks identified in **A**. Fraction of correlated genes in the hallmark is summarized at each *HL*_MRI_ interval of 0.0004. Significant different *HL*_MRI_ distributions are shown in each panel.

The data set of hypoxic fractions used for the analysis in Figure 5C was also included in a correlation analysis against HIF1A protein level assessed by immunohistochemistry (Fig. 7A). A strong correlation between HIF1A level and hypoxic fraction was found for a *HL*_MRI_ threshold of −0.21 (P=0.0021) (Fig. 7B), which was in the interval for mild hypoxia. This *HL*_MRI_ value was related to the cell line derived hypoxia activation level of 2.2% O_2_ (Fig. 5A), which is comparable to the findings for HIF1A stabilization in experimental studies (8,9). Moreover, the *HL*_MRI_ of −0.21 was outside the range for which a significant association to treatment outcome was found (Fig. 5C), consistent with results from survival analysis based on HIF1A protein (Fig. 7C). Taken together, by our imaging method it appeared possible to distinguish hypoxia levels with association to different biological processes like cancer hallmarks and HIF1A stabilization.

**Figure 7.**
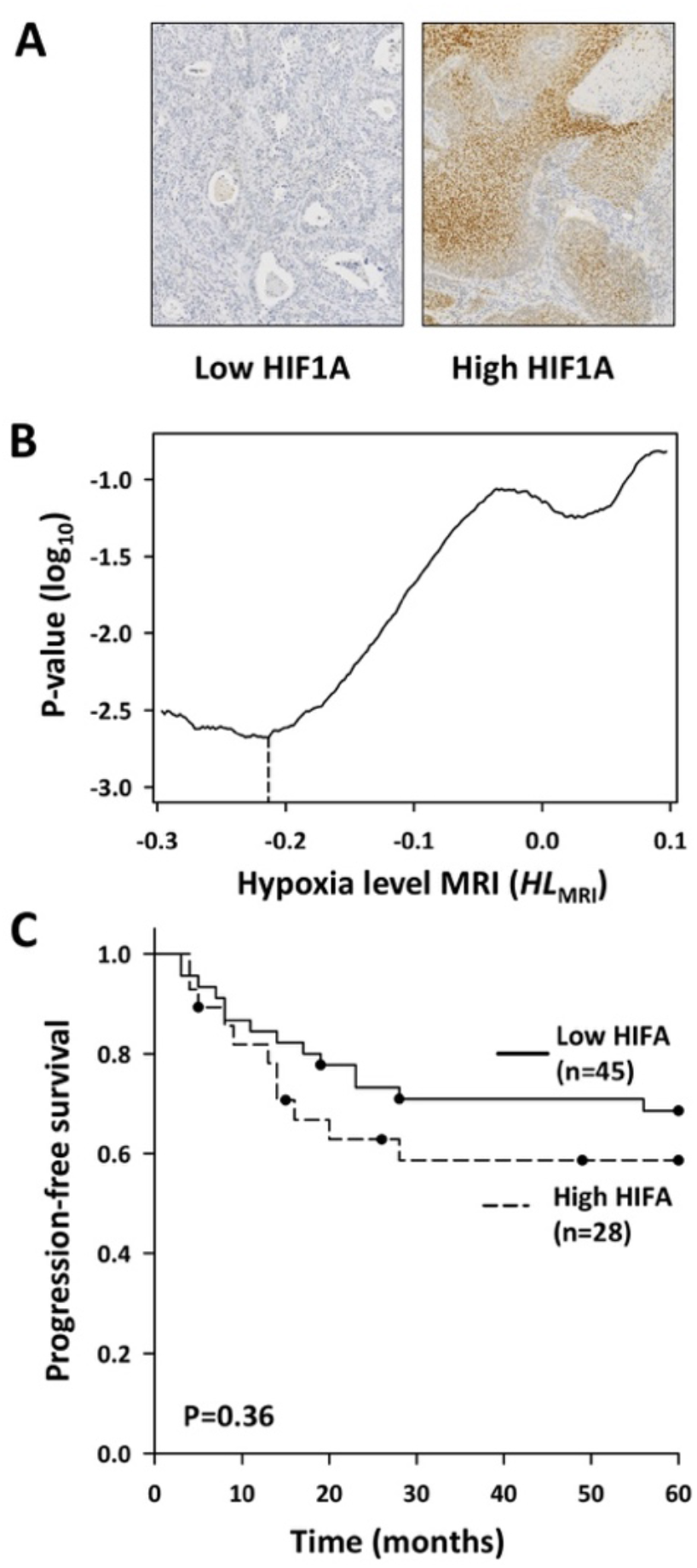
Hypoxia levels in patient tumors in relation to HIF1A protein level. **A**, Staining of HIF1A protein in a tumor with high (right) and low (left) protein level. **B**, P-value in correlation analysis of HIF1A protein level *versus* hypoxic fraction calcluated for increasing *HL*_MRI_ threshold (increasing severity level) in 73 patients, plotted as a function of *HL*_MRI_. Stippled line indicates *HL*_MRI_ for the strongest correlation (*HL*_MRI_=-0.21). **C**, Kaplan Meier curves for progression-free survival of 73 patients with low (solid line) and high (stippled line) level of HIFA protein. Patients were divided in two groups based on the pathology score, 0-3 and 4-5, to obtain approximately 1/3 in the high-risk and 2/3 in the low-risk group. *P*-value in log-rank test is shown.

## Discussion

We here present a method based on diagnostic MRI to visualize hypoxia levels in patient tumors. Previous imaging methods have focused solely on the presence of hypoxia without considering its severity (15). By utilizing the CSH-tool to combine multiparametric images, we obtained the weighted information of oxygen consumption and supply that visualized a continuous range of hypoxia levels. Although adding more information like cellular proliferation rate or blood oxyhemoglobin saturation may improve the technology, comparison of our results with direct measures of hypoxia levels by pimonidazole staining and indirect measures by gene expression showed strong correlations and validated the method. The hypoxia levels were found to differ in their association to treatment outcome and cancer hallmarks in cervical cancer, demonstrating that new understanding of how various levels affects tumor aggressiveness and biology can be achieved by our method. Our approach is easily applicable in the hospital’s diagnostic procedures, and is a step towards a better exploitation of MR images in the clinic.

Our algorithm to calculate hypoxia levels from MR images was validated in xenograft tumors by using pimonidazole staining intensity in histological sections as direct measure of hypoxia level. This approach was justified by our observation of a steady decrease in staining intensity away from necrosis. Binding of pimonidazole or other nitroimidazole compounds in cells or pieces of tumor tissue cultured *in vitro* under increasing oxygen concentrations has been shown to decrease in the same manner (24,26,27). Moreover, similar staining intensity gradients from necrosis have been quantified in tumor sections both by light and fluorescence microscopy (28,29). It is therefore likely that the intensity gradients in our histological sections reflected true differences in hypoxia levels. Further, by using large scale similarity analysis of hypoxic fractions obtained from MRI and pimonidazole staining, followed by correlation analysis of the corresponding levels, the linear range for reliable detection of hypoxia levels in xenograft tumors was obtained.

The algorithm was confirmed in patient tumors by using an indicator of hypoxia levels based on gene expression. We utilized that some genes are upregulated at specific levels because they primarily are involved in biological processes activated under these conditions (6). The hypoxia activation level has been assessed previously for the HIF1A protein as the oxygen concentration for half-maximal response in cell lines (8). The same strategy was applied on our gene expression data to construct a panel of indicator genes with different activation level. Strict criteria for gene selection, based on expression responses in two cell lines exposed to a range of oxygen concentrations and correlation analysis of expression and imaging data in patient tumors, revealed nine suitable indicator genes. Indeed, a strong linear relationship between the cell line derived hypoxia activation levels and *HL*_MRI_ was found, confirming that a continuous range of hypoxia levels could be visualized in patient tumors.

Caution should be taken to directly transfer the oxygen concentrations for activation of genes in cell lines to hypoxia levels in patient tumors, however, it would enable a rough comparison of our results with existing pO_2_ data of cervical cancer. The median MRI-defined hypoxia level for all tumors combined corresponded to a cell line derived level of 1.3% O_2_, which is within the range of 3-17 mmHg (approximately 0.4-2.2%) achieved by oxygen electrodes (14). Moreover, the strongest correlation to treatment outcome was found for a level corresponding to 0.7% O_2_ based on cell line data. This is highly consistent with most pO_2_ studies, reporting association to outcome for hypoxic fraction below 5 mmHg (approximately 0.7% O_2_) (14). Our approach therefore seemed to indicate hypoxia levels in accordance with oxygen electrode measurements, and to distinguish levels shown to be of prognostic significance in previous work.

Hypoxia levels associated with biological processes like cancer hallmarks and stabilization of HIF1A protein were identified by our method. At moderate hypoxia, which was the level most strongly correlated with treatment outcome, hallmarks like oxidative phosphorylation, targets of the MYC oncogene and G2/M checkpoint, appeared significant. This finding is consistent with our previous work where we identified a treatment resistant cervix tumor phenotype associated with the same hallmarks (30). In addition, this tumor phenotype appeared to have increased mitochondrial and proliferative activity (30). This implies that the stronger correlation of moderate hypoxia levels with poor outcome could be because hypoxic cells still have enough oxygen to proliferate under these conditions, in line with a hypothesis proposed by others (10,31). Stabilization of HIF1A protein, on the other hand, appeared significant at mild hypoxia levels, consistent with previous reports (6), and showed no correlation to outcome.

At severe hypoxia, the DNA repair hallmark appeared significant, consistent with studies showing activation of DNA damage response at extremely low oxygen concentrations (32). Our finding that inflammatory response and EMT were associated with such severe levels, on the other hand, is less well documented. It is tempting to speculate that this could be a consequence of lactate accumulation due to near complete vascular shut down in regions with severe hypoxia. Lactate is a key molecule in the inflammatory immune suppressive response in tumors (33,34), and such inflammatory environment is a strong inducer of EMT (35). Although these novel associations for the most severe levels need to be explored further in experimental work, the findings demonstrate a potential of our method to achieve better insight into the hypoxic tumor phenotype.

Our method to visualize hypoxia levels proposes a new application of routinously acquired DCE-MR images that may have implications for the diagnostic evaluation of patients. The finding that the CSH-tool could be used for this purpose, broadens the utility of the tool. This encourages investigations of hypoxia levels in other cancer types as well, by exploiting the MR technology already available at most hospitals. Our method provides a well needed opportunity to investigate the importance of individual hypoxia levels in tumor progression that eventually may lead to new and more efficient therapeutic options to combat hypoxia.

## Data Availability

Data is available at request. The gene expression data have been deposited to the GEO repository; GSE72723 (patient data), GSE147384 (cell line data).

## Acknowledgment

Technical assistance from D. Trinh, Department of Pathology, is highly appreciated.

